# Interpretable Machine Learning-based Decision Support for Prediction of Antibiotic Resistance for Complicated Urinary Tract Infections

**DOI:** 10.1101/2023.01.09.23284299

**Authors:** Jenny Yang, David W. Eyre, David A. Clifton

## Abstract

Urinary tract infections are one of the most common bacterial infections worldwide; however, increasing antimicrobial resistance in bacterial pathogens is making it challenging for clinicians to correctly prescribe patients appropriate antibiotics. In this study, we present four interpretable machine learning-based decision support systems for predicting antimicrobial resistance. Using electronic health record data from a large cohort of patients diagnosed with complicated UTIs, we demonstrate high predictability of antibiotic resistance across four antibiotics – nitrofurantoin, co-trimoxazole, ciprofloxacin, and levofloxacin. We additionally demonstrate the generalizability of our methods on a separate cohort of patients with uncomplicated UTIs. Through our study, we demonstrate that machine learning-based methods can help reduce the risk of non-susceptible treatment, facilitate rapid clinical intervention, enable more personalized approaches for treatment recommendation, while additionally allowing model interpretability that explains the basis for predictions.

## Introduction

Recent years have seen rapid increases in the prevalence of antimicrobial resistance in bacterial pathogens, which is threatening the efficacy of many antibiotic therapies, and ultimately leading to treatment failure (Ventola, 2015; Didelot & Pouwels, 2019; Yelin et al., 2019). Although new drugs are urgently needed, new antibiotic development is restricted by costs, limited government support, and regulatory requirements (Ventola, 2015; Didelot & Pouwels, 2019). Furthermore, antibiotic resistance leads to increased reliance on broad-spectrum therapies, which select for further resistance, exacerbating the issue at hand (Kanjilal et al., 2020; Yelin et al., 2019). To avoid these risks, it is critical for clinicians to correctly match available antibiotic therapies to the specific susceptibilities of bacterial pathogens, and ideally to do so at the point of starting empirical treatment before culture results are known, which may follow several days later. In this study, we present interpretable machine learning-based decision support systems for predicting antimicrobial resistance (AMR), which decreases the risk of non-susceptible and therefore, inactive treatment, and facilitates rapid effective clinical intervention. We demonstrate the utility of these systems for urinary tract infections (UTIs), where the problem of antibiotic resistance is of particular importance.

UTIs are one of the most common bacterial infections worldwide, affecting more than 150 million people each year (Flores-Mireles et al., 2015; Yelin et al., 2019). The pathogens that cause UTIs, including *Escherichia coli, Klebsiella pneumoniae, Proteus mirabilis, Enterococcus faecalis* and *Staphylococcus saprophyticus* (Pouwels et al., 2018; Flores-Mireles et al., 2015; Yelin et al., 2019) can often be carried asymptomatically and thus, are frequently exposed to antibiotics, including those intended for other infections (Didelot & Pouwels, 2019). This exposure, combined with high recurrence rates, often results in multidrug-resistant strains, with resistance rates of over 20% for commonly used drugs (Yelin et al., 2019). As treatment-outcome is associated with the infection-causing pathogen’s susceptibilities, clinicians are faced with the challenging task of correctly matching patients to an appropriate antibiotic (i.e., one that will effectively treat them). However, to provide rapid intervention, treatment is typically prescribed empirically (without any knowledge of which specific antibiotics the infection-causing pathogen are susceptible to), further compounding the risk of selecting inappropriate treatment (Yelin et al., 2019; Didelot & Pouwels, 2019).

Recent works have shown that machine learning-based algorithms, using electronic health record (EHR) data, including demographic information, prior antibiotic exposures, prior microbiology antibiotic susceptibility data, basic laboratory values, and comorbidities, can be used to predict antibiotic resistance in UTI infections. Analyzing six different antibiotics, Yelin et al. (2019) demonstrated that logistic regression and gradient-boosting decision trees could effectively improve the predictability of resistance (AUROC range 0.70-0.83), using demographics, microbiology sample history and antibiotic purchase history. Subsequently, they also found that the algorithm-suggested drug recommendations reduced the rate of mismatched treatments, both when using an unconstrained method (where the antibiotic with the lowest resistance probability was chosen) and a constrained method (where antibiotics were selected at the same frequency used by clinicians). Similarly, Kanjilal et al. (2020) used EHR data to predict the probability of antibiotic resistance for uncomplicated UTIs (i.e. those where the infection site was specified as urinary), achieving AUROCs between 0.56-0.64 across four different antibiotics. Despite relatively modest predictive performance, this still out-performed clinicians. In addition to predicting resistance, they also aimed to reduce the recommendation of broad-spectrum second-line therapies (e.g., fluoroquinolone antibiotics such as ciprofloxacin and levofloxacin, which have also been associated with serious adverse events in some patients). Using logistic regression and post-processing analysis, they found that their pipeline both reduced inappropriate antibiotic recommendations and achieved a 67% reduction in the recommendation of second-line agents, relative to clinicians. Although these studies found that logistic regression and gradient-boosting trees achieved the best results, neither investigated the effectiveness of neural network-based architectures.

Deep neural networks have notably been used for tasks involving image- and text-based data. However, it remains underexplored for tabular data, as ensemble-based decision trees (DTs) have typically achieved state-of-the-art success for such applications. One reason for this is that deep neural networks are overparametrized; and thus, the lack of inductive bias results in them failing to converge to optimal solutions on tabular decision manifolds (Arik & Pfister, 2021). Furthermore, a DT is highly interpretable, whereas a deep neural network is less straightforward to interpret, even commonly being referred to as a “black box” (Castelvecchi, 2016). This makes it difficult to implement neural networks for many real-world tasks, as model-interpretability is particularly important, especially for applications concerning clinical decision-making. However, there are many benefits to using neural networks, including improved performance on large datasets, and the ability to use transfer learning and self-/semi-supervised learning (Yang et al., 2022, Arik & Pfister, 2021). Moreover, with the advancements and increasing popularity of attention-based models (a type of sequence-to-sequence model), researchers have developed deep architectures capable of reasoning from features at each decision step, enabling model interpretability. One such model is the TabNet architecture (Arik & Pfister, 2021), which is uniquely tailored for interpretable learning from tabular data.

We aimed to expand on previous studies by 1) evaluating the utility of using ML-based prediction of antibiotic resistance for patients with complicated UTIs (namely, UTIs which are more severe in nature and/or occur in patients with anatomically abnormal urinary tracts or significant medical or surgical comorbidities [Neal, 2008]) and 2) demonstrating, comparing, and discussing the advantages of three types of interpretable machine learning methods, including a neural network-based model, which has not previously been used for predicting UTI resistance to antibiotics. We chose complicated UTIs, as these infections typically carry a higher risk of treatment failure due to prior antibiotic therapy, and are associated with more adverse outcomes with inactive treatment. These infections may also require longer courses of treatment and different antibiotics (Sabih & Leslie, 2022), emphasizing the necessity for novel intervention methods. Additionally, no previous works have focused on this cohort of patients for machine learning-based AMR tasks. Moreover, we specifically chose interpretable machine learning algorithms, as the ability to interpret and explain model predictions is important for clinical utility and facilitating use of machine learning models within routine care by clinicians. While antibiotic resistance for complicated UTIs was the motivating problem, the techniques introduced can be applied to many other applications.

## Methods

### Dataset, Features, and Preprocessing

We trained and tested our models using the AMR-UTI dataset (Oberst et al., 2020; Goldberger et al., 2000), which is a freely accessible dataset of over 80,000 patients with UTIs presenting between 2007 and 2016 at Massachusetts General Hospital (MGH) and Brigham & Women’s Hospital (BWH) (approved by the Institutional Review Board of Massachusetts General Hospital with a waived requirement for informed consent). Using this dataset, a previous study performed retrospective analyses on a subset of patients with uncomplicated UTI, consisting of 15,806 specimens (Kanjilal et al., 2020). This uncomplicated cohort was defined as specimens where the infection site was specified as urinary, and the following patient criteria were met: female between the ages of 18 to 55, no diagnosis indicating pregnancy in the past 90 days, no selected procedure (placement of a central venous catheter, mechanical ventilation, parenteral nutrition, hemodialysis, and any surgical procedure) in the past 90 days, no indication of pyelonephritis, and exactly one antibiotic of nitrofurantoin, co-trimoxazole, levofloxacin, or ciprofloxacin was prescribed. Thus, we instead focused our analyses on patients with potentially complicated UTI, totalling 101,096 specimens. This consisted of a broader cohort where patients did not satisfy the aforementioned requirements, including many patients with complex infections that may be treated with a range of antibiotics. We included all specimens that were tested for any one or combination of the local first-line agents – nitrofurantoin (NIT) or co-trimoxazole (SXT) – or second-line agents – ciprofloxacin (CIP) or levofloxacin (LVX).

To allow for direct comparison, we used a similar feature set and data filtering protocol as those used for the uncomplicated cohort. Each observation includes corresponding urine specimens which were sent to the clinical microbiology laboratory for assessment of AMR. Full de-identified feature sets include 1) the antimicrobial susceptibility profile (which we aim to predict), 2) previous specimen features useful for AMR prediction, and 3) basic patient features.

With respect to the antimicrobial susceptibility profile, the raw data received from the clinical microbiology laboratory included the identity of the infecting pathogen, alongside the results of susceptibility testing to various antibiotics. These were determined by minimum inhibitory concentration (MIC) and disk diffusion (DD) based methods, and the numerical results of these tests were transformed into categorical phenotypes using the published 2017 Clinical and Laboratory Standards Institute (CLSI) clinical breakpoints. This conversion resulted in three phenotypes: susceptible (S), intermediate (I), and resistant (R). The AMR-UTI dataset treated both intermediate and resistant phenotypes as resistant, which is typically in-line with what is done in clinical practice (Oberst et al., 2020). We adopt the same simplifying approach.

EHR data included patient demographic features such as age and ethnicity, prior antibiotic resistance, prior antibiotic exposures, prior infecting organisms, comorbidity diagnoses, where the specimen was collected (inpatient, outpatient, emergency room [ER], intensive care unit [ICU]), colonization pressure (rate of resistance to that agent within a specified location and time period), prior visits to skilled nursing facilities, infections at other sites (other than urinary), and prior procedures. Colonization pressure was computed as the proportion of all urinary specimens resistant to an antibiotic in the period ranging from 7 days before to 90 days before the date of specimen collection (for a given specimen), across 25 antibiotics. Resistance rates were recorded for three location hierarchies – specimens collected at the same floor/ward/clinic, specimens collected at the same hospital (MGH or BWH) and department type (inpatient, outpatient, ICU, ER), and all specimens collectively. Infections at other sites were included for those patients who had other specimens collected (on the same day as the urinary specimen) from other infection sites. Antibiotic exposures, prior resistances, prior infecting organisms, laboratory data, comorbidities, and prior hospitalizations were recorded for 14, 30, 90, and 180 days preceding specimen collection. These data do not include information on the dose or duration of antibiotic therapy, urinalysis results, drug allergies, or data for patient encounters outside of MGH and BWH. Empiric clinician prescriptions for patients diagnosed with complicated UTIs were not available in the dataset.

All categorical variables were one-hot encoded, totalling 787 features used for model development (a full list of features used can be found in Supplementary Table 2 [Section B of the Supplementary Material]). Missing values were already addressed within the dataset (as most features are binary, 1 indicates the presence of an observed element and 0 indicates that an element was not observed, including those cases where data is missing). Detailed documentation on data inclusion, exclusion, features, feature descriptions, and analytic protocols used for the AMR-UTI dataset can be found in the repository (https://physionet.org/content/antimicrobial-resistance-uti/1.0.0/).

Using the same features and preprocessing protocol, we additionally validate models using the cohort of patients with uncomplicated UTI (15,806 specimens). This is the same dataset used in a previous study (Kanjilal et al., 2020), allowing us to evaluate the generalizability of our models, as well as directly compare results to those from a previous benchmark.

To train and test our models, we used temporal evaluation, where models were trained on data from patients who submitted urine specimens between 2007 and 2013; and then tested on specimens submitted between 2014 and 2016. By temporally separating the data between training and test sets, we can emulate the real-world implementation of such a forecasting method for AMR. From the initial training data, we used 90% for model development, hyperparameter selection, and model training, and the remaining 10% for continuous validation and threshold adjustment of results. After successful model development and training, the held-out test set was used to evaluate the performance of the final models.

Patients in the training set cohort had a median age of 64 years (IQR 44-76), with 72.9% of patients self-identifying as white; the validation cohort also had a median age of 64 (44-76), with 73.6% self-identifying as white; and the test cohort had a median age of 64 (45-76), with 72.7% self-identifying as white. This differs from the uncomplicated UTI patient cohort presented in Kanjilal et al. (2020), who by definition were all female, and where the median age was 32 years (24-43), and 64.2% of patients self-identified as white (recall that the uncomplicated cohort specified an age range between 18-55). It should be noted that demographic information on the sex of patients in the complicated UTI cohort was not available in the dataset. Patients in the complicated UTI test set cohort presented more frequently in the emergency room (27.8% compared to 19.6% for the test set and training set cohorts, respectively). The prevalence of resistance to fluoroquinolones in the training, validation, and test set cohorts (with patient presentations between 2007-2016) was similar to national estimates reported in a cross-sectional survey in the United States in 2012 (Sanchez et al., 2016), which found that resistance was high among adults (11.8%) and elderly outpatients (29.1%) (compared to 21.6%-24.7% for training, validation, and test cohorts used in our study). For first-line therapies, the prevalence of resistance to SXT was similar to those reported in the study (22.3% and 26.8% for adults and older adults, respectively; compared to 22.3%-23.6% in our cohorts); however, the prevalence of resistance to NIT in our cohorts was higher (0.9% and 2.6% for adults and older adults, respectively; compared to 22.3%-22.5% in our cohorts). The majority of patients in our training, validation, and test cohorts had no prior drug resistance infections, recorded within the previous 90 days of the specimen sample (6.8%-6.9%, 6.5%-6.7%, 7.8%-9.0%, and 9.1%-9.7% for prior NIT, SXT, CIP, and LVX resistance, respectively, across training, validation, and test cohorts). A full summary of baseline characteristics for the training, validation, and test sets are presented in Supplementary Table 1 (Section B of the Supplementary Material).

### Machine Learning Architecture

We trained logistic regression, XGBoost, and TabNet models to predict the probability that a specimen would be resistant (i.e., non-susceptible) to NIT, SXT, CIP, or LVX. All models can handle tabular data consisting of both continuous and categorical features, and additionally, enable interpretability by quantifying the contributions of each feature to the trained model.

Logistic Regression (LR) is widely accepted in clinical decision-making, and additionally, has previously been shown to perform the best when evaluating uncomplicated UTI specimens, which were obtained using the same protocol as the complicated UTI cohort used in our study (Kanjilal et al., 2020). This makes it an appropriate benchmark for comparison to more complex models.

XGBoost is an optimized distributed gradient boosting library, based on decision trees (DTs), which has been found to achieve state-of-the-art results on many machine learning problems, especially those using structured or tabular datasets (as we use in our study). DT-based algorithms have also been shown to be effective at predicting AMR from clinical data (Yelin, 2019).

As previous AMR studies have not investigated the efficacy of neural network-based architectures, we trained TabNet architectures for each antibiotic. In addition to traditional supervised learning, we trained a separate set of TabNet models which have been pre-trained using self-supervised learning, specifically using unsupervised representation learning. Here, we train a decoder network to reconstruct the original tabular features from the encoded representations, through the task of predicting missing feature columns from the others. This ultimately results in an improved encoder model to be used during the main supervised learning task. Details about the TabNet architecture and the self-supervised method used can be found in the original TabNet publication (Arik & Pfister, 2021).

We individually trained LR, XGBoost, and TabNet models for each antibiotic; thus, training, validation, and test data slightly differed depending on whether a patient had susceptibility results for the antibiotic being tested. A summary of all training, validation, and test cohorts can be found in Table 1.

**Table 1:** Summary of the total number of specimens and non-susceptible cases in training, validation, and test set cohorts, for each antibiotic susceptibility prediction task.

|  | NIT | SXT | CIP | LVX |
| --- | --- | --- | --- | --- |
| **Training** |  |  |  |  |
| n, specimens | 58,972 | 53,865 | 57,631 | 61,586 |
| n, non-susceptible (%) | 13,925 (23.6%) | 13,851 (25.7%) | 13,495 (23.4%) | 15,123 (24.6%) |
| **Validation** |  |  |  |  |
| n, specimens | 6,553 | 5,986 | 6,404 | 6,843 |
| n, non-susceptible (%) | 1,514 (23.1%) | 1,561 (26.1%) | 1,492 (23.3%) | 1,711 (25.0%) |
| **Test** |  |  |  |  |
| n, specimens | 30,528 | 27,997 | 30,920 | 31,690 |
| n, non-susceptible (%) | 7,138 (23.4%) | 7,536 (26.9%) | 7,637 (24.7%) | 7,907 (25.0%) |

### Evaluation Metrics

To evaluate the trained models, sensitivity, specificity, area under the receiver operator characteristic curve (AUROC), and area under the precision recall curve (AUPRC) are reported, alongside 95% confidence intervals (CIs) based on 1000 bootstrapped samples taken from the test set.

### Hyperparameter Optimization and Threshold Adjustment

For each model developed, hyperparameter values were determined through standard five-fold cross-validation and grid search using respective training sets. This ensured that different combinations of hyperparameter values were evaluated on as much data as possible to provide the best estimate of model performance on unseen data and choose the optimal settings for model training. We chose the hyperparameter set based on the best AUPRC scores to account for the relative label imbalance in the dataset. Details on the hyperparameter values used in the final models can be found in Supplementary Table 3 (Section C of the Supplementary Material).

As the raw output of each classifier is a probability of class membership, a threshold is needed to map each specimen to a particular class label. For binary classification, the default threshold is typically 0.5 (values equal to or greater than 0.5 are mapped to one class and all other values are mapped to the other); however, this threshold can lead to poor performance, especially when the dataset used to train a model has a large class imbalance (Yang et al., 2022). This is seen in our training sets, as there are far fewer non-susceptible cases than susceptible ones, across all antibiotics. Thus, we used a grid search to adjust the decision boundary used for identifying non-susceptible specimens, to improve detection rates at the time of testing. We chose to optimize for balanced sensitivity and specificity to ensure that we can identify resistant samples (to avoid unsuccessful treatment), as well as ensure that samples which are susceptible get treated with the appropriate local first-line antibiotic (and avoid having to potentially use more antibiotics), respectively. The optimal thresholds were determined using the validation dataset, and were then applied to the results obtained on the held-out test set. Final threshold values used can be found in Supplementary Table 4 (Section C of the Supplementary Material).

## Results

After training models on cohorts of patients diagnosed with complicated UTI between 2007 and 2013, we temporally validated our models on patients diagnosed with complicated UTI between 2014 and 2016. Separate sets of models were trained to predict resistance for each of four antibiotics – NIT, SXT, CIP, and LVX. Overall, higher predictive performance was achieved by models developed for the second line antibiotics – CIP and LVX (mean AUROCs across all models of 0.800 [95% CIs ranged from 0.784-0.916] and 0.804 [0.786-0.810], respectively), than the first line antibiotics – NIT and SXT (0.674 [0.656-0.681] and 0.686 [0.660-0.707], respectively). For all antibiotics, XGBoost models achieved the best performances with respect to both AUROC and AUPRC (Table 2). LR and TabNet (without pre-training) models achieved the lowest AUROC and AUPRC scores, with non-overlapping CIs (except for the AUPRC CIs for the NIT model) when compared to XGBoost comparators, across all antibiotics, suggesting meaningful improvements were obtained through using the XGBoost architecture (p<0.001 across all antibiotics; p-value calculated by evaluating how many times XGBoost performs better than other models across 1000 pairs of iterations). However, when the TabNet models were pre-trained using a self-supervised method (TabNet^self^), AUROC and AUPRC scores improved across all antibiotics. Although overall predictive performance between TabNet^self^ and XGBoost models were similar, TabNet^self^ did not outperform the XGBoost comparators (p<0.001 for all antibiotics).

**Table 2:** Performance metrics for antibiotic resistance prediction for patients with complicated UTI. Results reported as AUROC, AUPRC, sensitivity, and specificity (alongside 95% confidence intervals) for NIT, SXT, CIP, and LVX resistance prediction.

| Antibiotic | Model | AUROC | AUPRC | Sensitivity | Specificity |
| --- | --- | --- | --- | --- | --- |
| NIT | LR | 0.662(0.656-0.668) | 0.381(0.371-0.390) | 0.623(0.614-0.633) | 0.619(0.614-0.624) |
|  | XGBoost | **0.686(0.681-0.693)** | **0.411(0.401-0.421)** | 0.673(0.664-0.682) | 0.590(0.585-0.595) |
|  | TabNet | 0.670(0.664-0.677) | 0.393(0.383-0.403) | 0.674(0.664-0.683) | 0.565(0.559-0.570) |
|  | TabNet<sup>self</sup> | **0.676(0.670-0.682)** | **0.396(0.386-0.405)** | 0.634(0.625-0.644) | 0.622(0.617-0.627) |
| SXT | LR | 0.666(0.660-0.673) | 0.467(0.458-0.478) | 0.568(0.559-0.577) | 0.674(0.669-0.680) |
|  | XGBoost | **0.701(0.695-0.707)** | **0.524(0.514-0.534)** | 0.660(0.651-0.669) | 0.618(0.612-0.623) |
|  | TabNet | 0.685(0.678-0.691) | 0.497(0.487-0.508) | 0.629(0.620-0.639) | 0.635(0.630-0.641) |
|  | TabNet<sup>self</sup> | **0.693(0.687-0.699)** | **0.503(0.492-0.513)** | 0.637(0.628-0.646) | 0.641(0.635-0.646) |
| CIP | LR | 0.789(0.784-0.794) | 0.590(0.580-0.599) | 0.571(0.561-0.580) | 0.855(0.851-0.859) |
|  | XGBoost | **0.811(0.806-0.816)** | **0.617(0.608-0.627)** | 0.696(0.687-0.705) | 0.783(0.778-0.788) |
|  | TabNet | 0.798(0.793-0.802) | 0.576(0.566-0.586) | 0.707(0.699-0.716) | 0.755(0.750-0.759) |
|  | TabNet<sup>self</sup> | **0.800(0.796-0.805)** | **0.584(0.575-0.595)** | 0.726(0.718-0.734) | 0.737(0.733-0.742) |
| LVX | LR | 0.791(0.786-0.796) | 0.592(0.582-0.602) | 0.632(0.623-0.641) | 0.809(0.805-0.813) |
|  | XGBoost | **0.814(0.810-0.819)** | **0.624(0.614-0.634)** | 0.710(0.702-0.719) | 0.769(0.765-0.773) |
|  | TabNet | 0.803(0.798-0.808) | 0.597(0.587-0.608) | 0.725(0.718-0.734) | 0.737(0.732-0.741) |
|  | TabNet<sup>self</sup> | **0.808(0.803-0.813)** | **0.606(0.597-0.617)** | 0.713(0.705-0.721) | 0.764(0.760-0.769) |

To evaluate the generalizability of our models, we additionally performed validation on an independent cohort of patients with uncomplicated UTI specimens. We used the trained XGBoost and TabNet^self^ models, as these achieved the best and second-best scores during temporal validation on the complicated UTI specimens. We present results for all specimens (n=15,608), as well as results for a smaller subset (n=3,941) which is equivalent to the test set evaluated in Kanjilal et al. (2020), allowing for direct comparison. For all antibiotics, AUROC and AUPRC are lower for the uncomplicated cohort than the complicated cohort; however, they are comparable to those reported in the previous study, despite the previous study being specifically trained on uncomplicated UTI, and this study being trained on complicated UTI (Table 3).

**Table 3:** Performance metrics for antibiotic resistance prediction for patients with uncomplicated UTI. Results reported as AUROC, AUPRC, sensitivity, and specificity (alongside 95% confidence intervals) for NIT, SXT, CIP, and LVX resistance prediction.

|  |  | TabNet<sup>self</sup> |  | XGBoost |  | Kanjilal et al. (2020) |
| --- | --- | --- | --- | --- | --- | --- |
|  |  | AUROC | AUPRC | AUROC | AUPRC | AUROC |
| NIT | All | 0.575(0.563-0.587) | 0.172(0.159-0.185) | 0.593(0.580-0.605) | 0.186(0.173-0.200) |  |
|  | Test | 0.543(0.517-0.566) | 0.145(0.128-0.169) | 0.559(0.534-0.584) | 0.162(0.142-0.187) | 0.56(0.53-0.59) |
| SXT | All | 0.603(0.594-0.613) | 0.301(0.289-0.315) | 0.612(0.603-0.621) | 0.318(0.305-0.331) |  |
|  | Test | 0.591(0.571-0.61) | 0.292(0.268-0.32) | 0.589(0.57-0.608) | 0.294(0.268-0.322) | 0.59(0.57-0.62) |
| CIP | All | 0.670(0.651-0.688) | 0.249(0.225-0.276) | 0.676(0.659-0.694) | 0.254(0.23-0.281) |  |
|  | Test | 0.646(0.611-0.679) | 0.244(0.199-0.294) | 0.639(0.606-0.673) | 0.245(0.202-0.294) | 0.64(0.60-0.68) |
| LVX | All | 0.662(0.644-0.678) | 0.228(0.204-0.255) | 0.667(0.649-0.685) | 0.244(0.22-0.273) |  |
|  | Test | 0.639(0.604-0.671) | 0.256(0.215-0.304) | 0.623(0.586-0.657) | 0.266(0.221-0.314) | 0.64(0.60-0.68) |

Overall, the results show promise that model-assigned probabilities of antibiotic resistance can differentiate complicated UTI specimens resistant to one antibiotic and susceptible to another at the single-patient level. Additionally, we found that the trained models can be generalized to uncomplicated UTI specimens, thus motivating further development of algorithmic decision-support for antibiotic recommendations.

Beyond solely classifying samples, all models can provide information on which features were most important for determining resistance (in the form of coefficients for logistic regression, and importance scores for TabNet and XGBoost models). For all models, prior antibiotic resistance and prior antibiotic exposure, across different time frames, were generally found to be the most important features in predicting resistance to each antibiotic. This included previous use of common antibiotics for UTI treatment (both the outcome antibiotics considered in our study, as well as other antibiotics) such as fluoroquolines (e.g. CIP and LVX), cephalosporins (e.g. cefepime, ceftriaxone, cefpodoxime), and penicillins (e.g. amoxicillin).

Similarly, previous UTI history (i.e. if any – susceptible or non-susceptible – isolates of infecting pathogens, such as *E*.*coli*, were found within previous patient specimens), was found to be predictive of resistance. For the second-line antibiotics (such as CIP and LVX), resistance to one was predictive of resistance to the other, which is expected, as both antibiotics belong to the same family of antibacterial agents. Additionally, comorbidities, including those categorized as paralysis and renal, were ranked highly across all antibiotics and models. Previous stays in a long-term care facility (skilled nursing facility) and whether a patient had undergone a surgical procedure were also considered as highly predictive factors. A full summary of the top 30 features used in prediction for each model, alongside their importance scores, can be found in Section D of the Supplementary Material.

Finally, we grouped features into sets that corresponded to general risk factor domains that were found to be associated with resistance. Using the XGBoost model architecture (as this achieved the best performances on the test set), we evaluated the decrease in predictive performance when a particular feature set was left out of training (Figures 1 and 2 for AUROC and AUPRC scores, respectively). In general, prior antibiotic resistance was found to be the most important feature set in predicting antibiotic resistance. When left out, AUPRC decreased by 0.0199 (0.0142-0.0265), 0.0877 (0.0800-0.0947), 0.0695 (0.0623-0.0759), and 0.0631 (0.0566-0.0692), for NIT, SXT, CIP, and LVX, respectively (for all antibiotics, decrease in AUPRC was found to be significant when compared to XGBoost models that were trained with all feature sets; p<0.001, determined using 1000 bootstrap samples). Prior antibiotic exposure was also found to be an important feature set, as AUPRC decreased by 0.0089 (0.0035-0.0142), 0.0401 (0.0035-0.0142), 0.0383 (0.0327-0.0441), and 0.0414 (0.0352-0.0472), for NIT, SXT, CIP, and LVX, respectively (p=0.001 for NIT, and p<0.001 for SXT, CIP, and LVX models). This aligns with the feature rankings obtained through the importance scores/coefficients quantified by each trained model. Although the absence of prior infecting organism features (i.e. prior UTI history) in training decreased predictive performance (AUPRC scores decreased by 0.0049 [0.0019-0.0072], 0.0015 [−0.0019-0.0052], 0.0038 [0.000-0.0079], and 0.0034 [−0.0003-0.0068] for NIT, SXT, CIP, and LVX, respectively), changes in AUPRC and AUROC scores were not generally found to be statistically significant (p=0.001, 0.403, 0.059, 0.062 for NIT, SXT, CIP, and LVX models, respectively). Similar patterns were found for AUROC scores. Full numerical results can be found in Supplementary Tables 9, 10, and 11 (Section E in the Supplementary Material).

**Figure 1:**
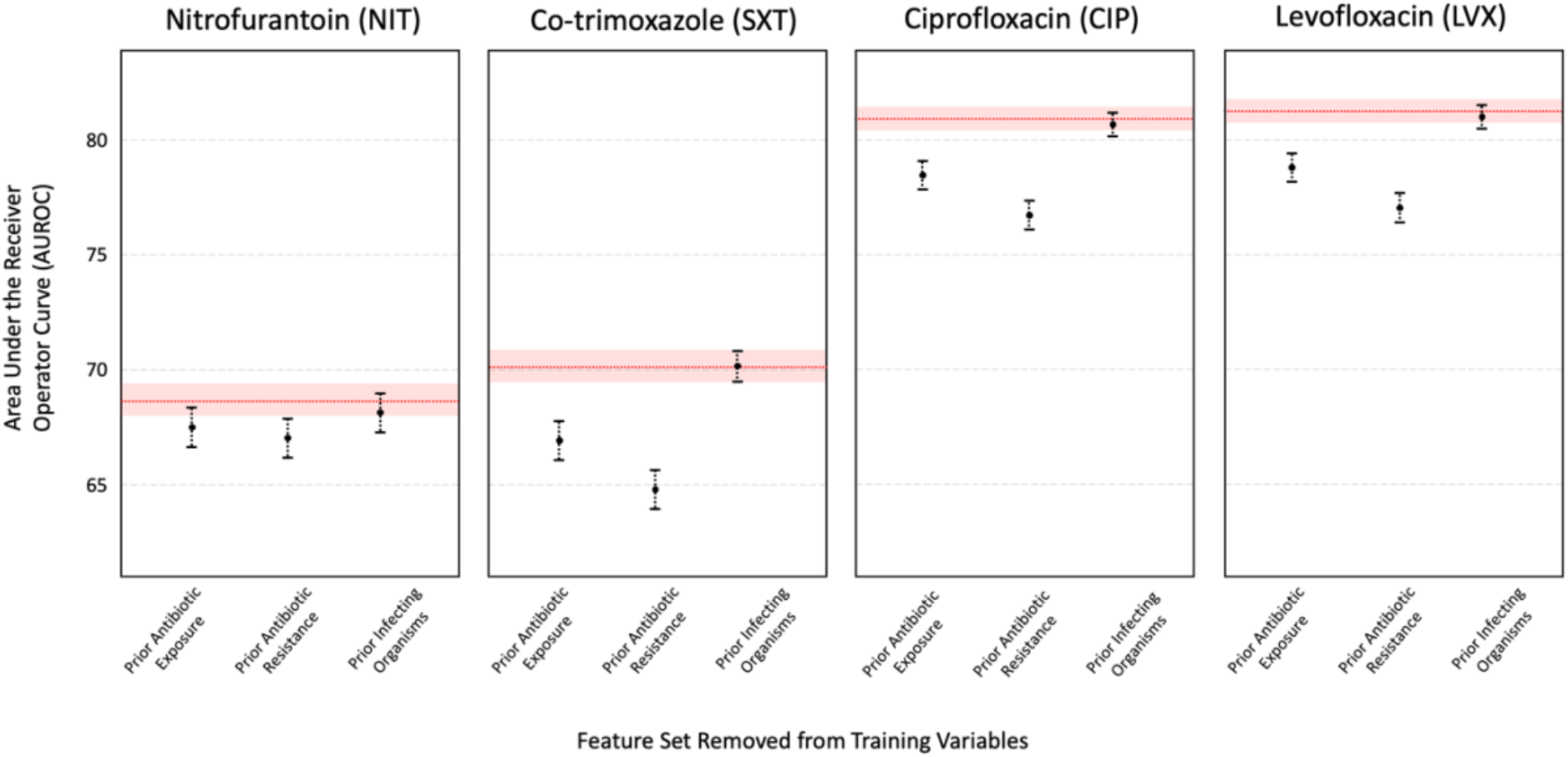
AUROC of XGBoost models trained without the feature set labeled on the x-axis, with error bars representing 95% CIs. The red line depicts the AUROC for the model trained on all features, with the red shaded region representing 95% CIs.

**Figure 2:**
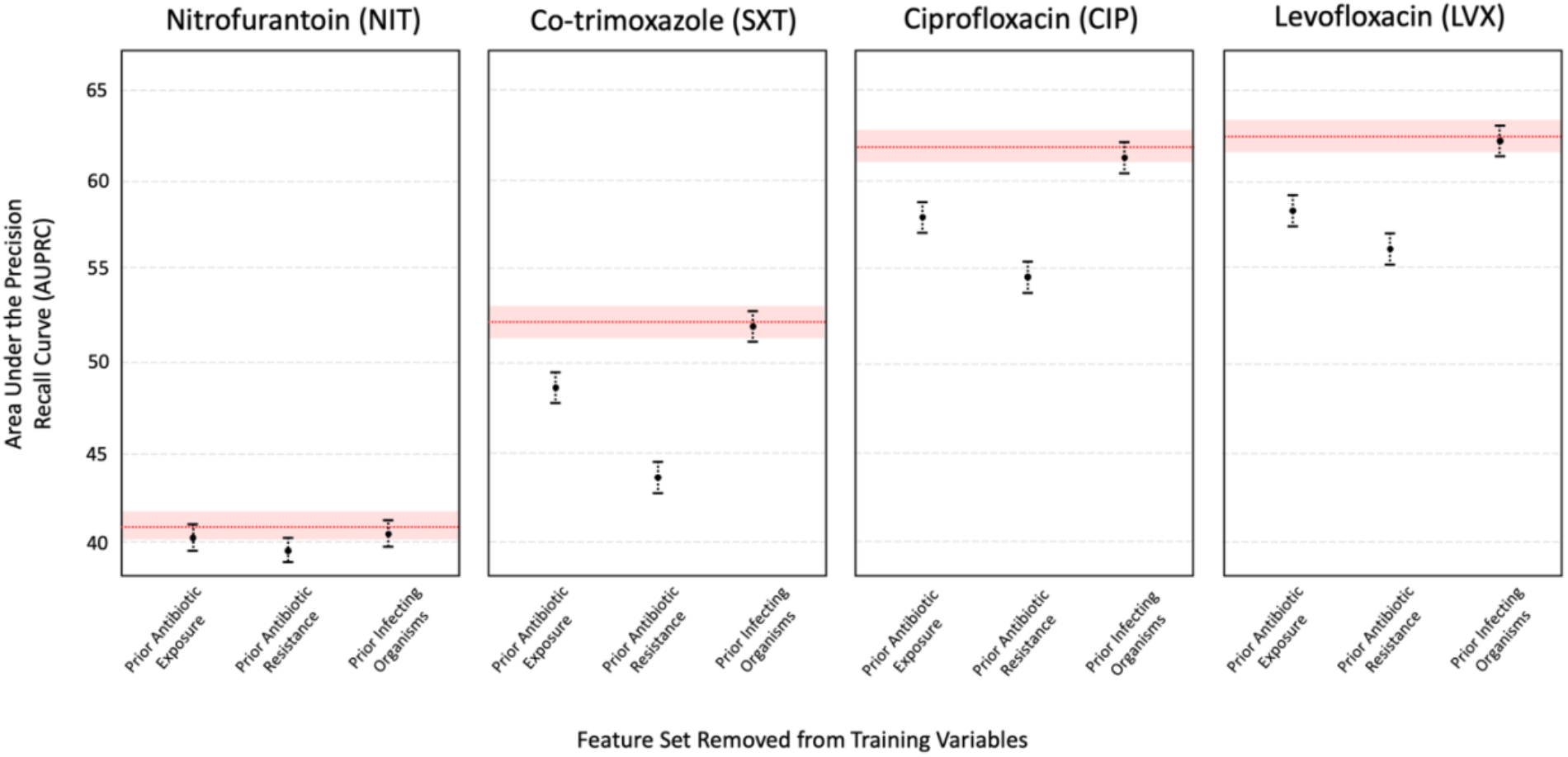
AUPRC of XGBoost models trained without the feature set labeled on the x-axis, with error bars representing 95% CIs. The red line depicts the AUPRC for the model trained on all features, with the red shaded region representing 95% CIs.

## Conclusion and Discussion

Through analyzing electronic health records from a large cohort of patients, we demonstrated high predictability of antibiotic resistance in complicated UTIs, using ML-based decision support methods. In addition to temporal validation (which emulates the real-world implementation of such algorithms), we also demonstrated the generalizability of our methods on a separate cohort of patients with uncomplicated UTI. Through evaluating four interpretable machine learning methods, we hope to encourage increased development and adoption of machine learning models within routine care, to both reduce the chances of mismatched treatment and to enable more personalized approaches for treatment recommendation.

We found that both XGBoost and TabNet models outperformed logistic regression, suggesting that there may be non-linear trends and interactions that cannot be sufficiently expressed as a linear combination of the input features. The best performances were achieved using XGBoost, which may be a result of its ensemble architecture, whereby the predictions of multiple models are combined, improving generalization error. However, TabNet, when combined with self-supervised learning, also achieved high performance (within CIs of the respective XGBoost comparators). XGBoost may have also outperformed TabNet because of the nature of the training data itself. Decision tree-based methods (such as XGBoost) have been found to perform better on discrete/categorical features compared to neural networks (which is the basis of TabNet); and thus, since our feature set is heterogeneous (containing both continuous and discrete features), XGBoost may have outperformed TabNet. However, there are advantages to using a neural network-based architecture (such as TabNet) including 1) it can be used in combination with transfer learning and self-supervised learning (which we demonstrated here), whereas tree-based algorithms, such as XGBoost, typically depend on the availability of the entire dataset, making transfer learning infeasible; and 2) it can be used for image recognition tasks, as well as natural language problems, which XGBoost is not appropriate for.

The ability of a TabNet model to use transfer learning may be of particular importance in a clinical context to allow predictions to be updated over time, or to be transferred to new hospital locations with variations in the prevalence of antimicrobial resistance or variation in clinical practice and patient characteristics. Here, through the acquisition of more data, the weights of a neural network base model can be finetuned, rather than fully retrained, as typically required by tree-based algorithms such as XGBoost.

We also found that we achieved better performance (with respect to AUROC) on the complicated UTI cohort than the uncomplicated UTI cohort. This may be because of greater hospital exposure (and related factors) in the complicated UTI cohort, making it easier to predict antibiotic susceptibility, compared to the uncomplicated cohort. Although we trained our models on the complicated UTI cohort, we still achieved comparable AUROC scores when validating on the uncomplicated cohort, as a previous study which trained and tested models using exclusively data from an uncomplicated UTI cohort. This may be due to the greater amount of data available for training, as machine learning models (and particularly, neural networks) typically need a large amount of training data to achieve generalizability. Additionally, this previous study outperformed clinicians in prescribing effective antibiotic therapies, and since we achieved similar performance to this earlier study, we would also expect our predictions to outperform empiric clinical prescriptions for the complicated UTI cohort (which – at the time of our study – we did not have available for us to evaluate).

Prior antibiotic resistance and antibiotic exposure (of both the outcome antibiotic, and others) was found to be highly predictive of resistance across all antibiotics. This is expected as antibiotic resistance has been found to be associated with previous UTI occurrences and their resistances (Ikram et al., 2015; Yelin et al., 2019; MacFadden et al., 2014). These features were ranked highly across multiple time frames proceeding specimen collection, suggesting both short- and long-term associations with resistance. In our investigations, times were binned; however, future studies may benefit from keeping a higher degree of granularity, as well as using models more suitable for time-series analysis/forecasting (e.g. convolutional neural network, long short-term memory network), to better capture temporal associations. Other antibiotic exposures (other than antibiotic being tested) were also ranked highly amongst the models. This is consistent with previous studies (Odoki et al., 2020; Chen et al., 2012; Bader et al., 2017), where a specific antibiotic exposure was found to both directly select for strains resistant to it, as well as indirectly select for resistance to other antibiotics (e.g. through common co-occurrence).

In addition to antibiotic-related features, comorbidities, including those categorized as paralysis and renal, were also commonly ranked as being important for determining resistance. These have previously been found to be associated with UTIs – patients with prior kidney diseases are at higher risk of developing UTIs (Scherberich et al., 2021); and patients with paralysis may have had a catheter-associated UTI (CAUTI), as catheters have been found to be a common cause of healthcare-associated UTIs (Bader et al., 2017; Ikram et al., 2015; Magill et al., 2014). Both of these factors can lead to recurrent UTIs; and thus, lead to antibiotic resistance due to prior exposure/use. This may also reflect why stays in a long-term care facility or undergoing a surgical procedure were also ranked as highly predictive, as patients may require the use of a catheter (Bader et al., 2017; Lin et al., 2021). Additionally, patients undergoing long surgical procedures may have postoperative urinary retention, which can also lead to a UTI (Pertsch et al., 2021).

We used threshold adjustment to determine a final susceptibility label, optimizing for a balanced sensitivity and specificity. This technique is especially useful when there are large imbalances in the training data (which we had in our training sets). However, it has previously been shown that the output of a model can be biased on the specific dataset it was trained on; thus, the optimal threshold can also be biased on the particular dataset used for derivation (Yang et al. 2022). Thus, the threshold used for one dataset (e.g. from a particular hospital, cohort or temporal range), may not be suitable for another dataset with independent distributions. Thus, determining an optimal decision threshold should be further investigated, as it directly affects model performance (namely, through the shift in true positive/true negative rates). This is also of particular importance for clinical tasks, as achieving consistent sensitivity/specificity scores during testing is desirable, as varying sensitivities/specificities across testing cohorts can make it difficult for clinicians to rely on the performance characteristic of a model (Yang et al, 2022). As real-world implementation of such methods need to consider temporal testing, future experiments can consider incrementally calibrating thresholds during deployment (to correspond to real-time distributions), in order to standardise predictive performance. Developers can also consider balancing data during the data pre-processing stage (before model training), as to relieve data-imbalance issues (and avoid threshold adjustment); however, we chose not to opt for this method, as we wanted to consider true prevalence rates during model development.

With respect to threshold adjustment, the trade-off between sensitivity and specificity should be carefully considered and tailored towards each task-at-hand. For our purposes, we optimized for balanced sensitivity and specificity, as we wanted to both avoid unsuccessful treatment, as well as ensure susceptible samples get treated with the appropriate local first-line antibiotic. However, depending on the prediction task, it may be more important to optimize for either sensitivity (e.g. in tasks where a missed diagnosis can lead to adverse events) or specificity (e.g. in tasks where a low specificity can lead to an unnecessary increase in resource use and costs, overburdening hospitals).

Because each patient can be resistant to multiple antibiotics, future studies can consider training one multilabel classifier/learning combinations of resistances, rather than multiple binary classifiers (as we’ve demonstrated here). This may be beneficial, as resistances to one antibiotic can affect the resistance to others; and thus, a single model that considers all antibiotics can account for the fact that patients can have multiple resistances. Additionally, for tasks where very large models need to be used, training multiple binary models can overwhelm computing power. However, multilabel tasks often require more data to confidently differentiate between all classes, especially for challenging clinical tasks.

We also appreciate that the probability of antibiotic susceptibility is a useful measurement, as opposed to thresholding to a binary label. We used a binary classification to align with the CLSI clinical breakpoints used in the AMR-UTI dataset; however, probability can also be used as a final output for tasks where appropriate.

Finally, we acknowledge that these findings are specific to this patient cohort in particular (data from MGH and BWH during 2007-2016); and thus, results may differ for other domains and datasets (varying in patient demographic distribution, type, size, features, etc.) and output type (e.g. regression). However, as neural network-based models enable transfer learning and finetuning, domain adaptation can be an interesting area to explore in future studies.

## Data Availability

The AMR-UTI data can be downloaded from: https://physionet.org/content/antimicrobial-resistance-uti/1.0.0/.

## Contributions

JY conceived and designed the study. JY preprocessed the data, wrote the code, performed the analyses, and wrote the manuscript. All authors revised the manuscript.

## Acknowledgements

We express our sincere thanks to all patients and staff across Massachusetts General Hospital (MGH) and Brigham & Women’s Hospital (BWH). We additionally express our gratitude to the MIT Clinical ML group for obtaining the data and making it freely accessible to other researchers.

## Ethics

The AMR-UTI dataset is a publicly-available, anonymized dataset, approved by the Institutional Review Board (IRB) of Massachusetts General Hospital with a waived requirement for informed consent.

## Data and Code Availability

The AMR-UTI data can be downloaded from: https://physionet.org/content/antimicrobial-resistance-uti/1.0.0/. Code will be available alongside publication.

