## Supplementary Material for "Interpretable Machine Learning-based Decision Support for Prediction of Antibiotic Resistance for Complicated Urinary Tract Infections"

#### A Software:

Models were implemented using Python (v3.6.9). XGBoost baseline models were implemented using the XGBoost library (v1.3.3). Logistic regression models were implemented using sklearn (v0.24.1). TabNet models were implemented using the pytorch-tabnet package (4.0) and torch (v1.7.0) All models were run using an Intel Xeon E-2146G Processor (CPU: 6 cores, 4.50 GHz max frequency).

#### B Data:

Table 1: Demographics, location of specimen collection (may have more than one location), and microbiologic characteristics for patients with complicated UTI. P-values are calculated using the Kruskal-Wallis Test. *percentage calculated relative to the entire dataset (original dataset prior to any pre-processing, including specimens where antibiotic result was not available – values differ slightly when separated into NIT, SXT, CIP, LVX cohorts)

| | Training (2007-2013) | Validation (2007-2013) | Test (2014-2016) | p |
|---|---|---|---|---|
| n (specimens) | 62,187 | 6,910 | 31,999 | |
| Age, mean (SD) | 58.6 (22.5) | 58.5 (22.4) | 59.2 (21.8) | 0.004 |
| Race, n (%) | | | | |
| White | 45,361 (72.9%) | 5,086 (73.6%) | 23,266 (72.7%) | 0.444 |
| Non-white | 16,826 (27.1%) | 1,824 (26.4%) | 8,733 (27.3) | |
| Location, n (%) | | | | |
| Outpatient | 31,262 (50.3%) | 3,439 (49.8%) | 15,528 (48.5%) | <0.001 |
| Emergency Room | 12,206 (19.6%) | 1,344 (19.5%) | 8,900 (27.8%) | <0.001 |
| General Inpatient | 16,141 (26.0%) | 1,825 (26.4%) | 6,606 (20.6%) | <0.001 |
| Intensive Care Unit | 3,417 (5.5%) | 3,88 (5.6%) | 1,311 (4.1%) | <0.001 |
| Current Resistance*, n | | | | |
| (%) | 13,883 (22.3%) | 1,556 (22.5%) | 7,138 (22.3%) | 0.565 |
| NIT | 13,858 (22.3%) | 1,554 (22.5%) | 7,536 (23.6%) | <0.001 |
| SXT | 13,453 (21.6%) | 1,534 (22.2%) | 7,637 (23.9%) | <0.001 |
| CIP | 15,130 (24.3%) | 1,704 (24.7%) | 7,907 (24.7%) | 0.205 |
| LVX | | | | |
| Prior Resistance (in past 90 days)*, n (%) | | | | |
| NIT | 4,269 (6.9%) | 467 (6.8%) | 2,207 (6.9%) | 0.967 |
| SXT | 4,133 (6.6%) | 448 (6.5%) | 2,159 (6.7%) | 0.439 |
| CIP | 5,082 (8.2%) | 536 (7.8%) | 2,864 (9.0%) | <0.001 |
| LVX | 6,059 (9.7%) | 626 (9.1%) | 2,942 (9.3%) | 0.004 |

Table 2: Full list of features included in training.

| Patient Demographics/ Hospital department type/ Prior visits to skilled nursing facilities (recorded at <7, <14, <30, <90 days prior to specimen collection) | Prior Resistances (recorded at <14, <30, <90, <180, and all days prior to specimen collection) | Prior Infecting Organisms (recorded at <14, <30, <90, and <180 days prior to specimen collection) | Prior Antibiotic Exposure (recorded at <7, <14, <30, <90, <180, and all days prior to specimen collection) | Comorbidities (recorded at <7, <14, <30, <90, <180 days prior to specimen collection) | Prior procedures (<180 days prior to specimen collection) | Other infection sites | Colonization pressure (recorded for granular, higher, and overall levels) |
|---|---|---|---|---|---|---|---|
| 'demographics – age',<br>'demographics – is_white',<br>'hosp ward - ER',<br>'hosp ward - ICU',<br>'hosp ward - IP',<br>'hosp ward - OP',<br>'custom - nursing home' | 'micro - prev resistance LVX',<br>'micro - prev resistance AMP',<br>'micro - prev resistance CFZ',<br>'micro - prev resistance CIP,<br>'micro - prev resistance GEN,<br>'micro - prev resistance PIP',<br>'micro - prev resistance ATM,<br>'micro - prev resistance IPM,<br>'micro - prev resistance TIC,<br>'micro - prev resistance TOB,<br>'micro - prev resistance ERY,<br>'micro - prev resistance OXA',<br>'micro - prev resistance PEN',<br>'micro - prev resistance CLI,<br>'micro - prev resistance NAL,<br>'micro - prev resistance DOX,<br>'micro - prev resistance QUD',<br>'micro - prev resistance RIF,<br>'micro - prev resistance TET,<br>'micro - prev resistance AMC,<br>'micro - prev resistance AMK,<br>'micro - prev resistance CAZ,<br>'micro - prev resistance CHL,<br>'micro - prev resistance CRO,<br>'micro - prev resistance CTT',<br>'micro - prev resistance FEP,<br>'micro - prev resistance NIT',<br>'micro - prev resistance SXT,<br>'micro - prev resistance VAN,<br>'micro - prev resistance TZP,<br>'micro - prev resistance ERT',<br>'micro - prev resistance LZD,<br>'micro - prev resistance SAM',<br>'micro - prev resistance STRS',<br>'micro - prev resistance GENS',<br>'micro - prev resistance MXF',<br>'micro - prev resistance FOX, | 'micro - prev organism Pseudomonas',<br>'micro - prev organism Escherichia ',<br>'micro - prev organism Staphylococcus ',<br>'micro - prev organism Enterococcus ',<br>'micro - prev organism Enterobacter ',<br>'micro - prev organism Providencia ',<br>'micro - prev organism Staph_coag_neg ',<br>'micro - prev organism Streptococcus ',<br>'micro - prev organism Acinetobacter ',<br>'micro - prev organism Citrobacter ',<br>'micro - prev organism Klebsiella ',<br>'micro - prev organism Proteus ',<br>'micro - prev organism Serratia ',<br>'micro - prev organism Morganella ' | 'medication - levofloxacin',<br>'medication - vancomycin',<br>'medication - ciprofloxacin',<br>'medication - azithromycin',<br>'medication - trimethoprim/sulfamethoxazole',<br>'medication - nitrofurantoin'<br>,<br>'medication - tetracycline',<br>'medication - moxifloxacin',<br>'medication - fluconazole',<br>'medication - amoxicillin',<br>'medication - cephalexin',<br>'medication - cefpodoxime',<br>'medication - doxycycline',<br>'medication - cefixime',<br>'medication - ceftriaxone',<br>'medication - amoxicillin/clavulanate',<br>'medication - dicloxacillin',<br>'medication - meropenem',<br>'medication - cefepime',<br>'medication - micafungin',<br>'medication - metronidazole'<br>,<br>'medication - clindamycin',<br>'medication - ofloxacin',<br>'medication - gentamicin',<br>'medication - minocycline',<br>'medication - clarithromycin<br>',<br>'medication - amikacin',<br>'medication - ertapenem',<br>'medication - cefuroxime',<br>'medication - cefazolin',<br>'medication - trimethoprim',<br>'medication - fosfomycin',<br>'medication - ampicillin/sulbactam',<br>'medication - penicillin',<br>'medication - aztreonam',<br>'medication - cefotetan',<br>'medication - rifampin',<br>'medication - nafcillin',<br>'medication - ceftazidime',<br>'medication - linezolid',<br>'medication - erythromycin',<br>'medication - cefaclor',<br>'medication - caspofungin',<br>'medication - cefoxitin',<br>'medication - polymyxin_b',<br>'medication - amphotericin_b',<br>'medication - cefadroxil',<br>'medication - cefprozil',<br>'medication - amoxicillin_clarithromycin'<br>'ab subtype - azole',<br>'ab subtype - penicillins',<br>'ab subtype - cephalosporin_gen1',<br>'ab subtype - cephalosporin_gen3',<br>'ab subtype - beta_lactam_combo',<br>'ab subtype - antistaphylococcal',<br>'ab subtype - carbapenem',<br>'ab subtype - cephalosporin_gen4',<br>'ab subtype - echinocandin',<br>'ab subtype - cephalosporin_gen2',<br>'ab subtype - polyene']<br>'ab class - fluoroquinolone',<br>'ab class - glycopeptides',<br>'ab class - macrolide_lincosamide',<br>'ab class - folate_inhibitor',<br>'ab class - nitrofuran',<br>'ab class - tetracycline',<br>'ab class - antifungal',<br>'ab class - beta_lactam',<br>'ab class - nitroimidazole',<br>'ab class - aminoglycoside',<br>'ab class - fosfomycin',<br>'ab class - monobactam',<br>'ab class - ansamycin',<br>'ab class - oxazolidinones',<br>'ab class - polymyxin',<br>'ab class - mixed' | 'comorbidity - Arrhythmia',<br>'comorbidity - HTN',<br>'comorbidity - Hypothyroid',<br>'comorbidity - Pulmonary',<br>'comorbidity - Depression',<br>'comorbidity - CHF',<br>'comorbidity - FluidsLytes',<br>'comorbidity - Lymphoma',<br>'comorbidity - Obesity',<br>'comorbidity - DM',<br>'comorbidity - PVD',<br>'comorbidity - Rheumatic',<br>'comorbidity - Anemia',<br>'comorbidity - BloodLoss',<br>'comorbidity - Renal',<br>'comorbidity - Paralysis',<br>'comorbidity - NeuroOther',<br>'comorbidity - Valvular',<br>'comorbidity - HTNcx',<br>'comorbidity - Coagulopathy',<br>'comorbidity - DMcx',<br>'comorbidity - Alcohol',<br>'comorbidity - Tumor',<br>'comorbidity - Psychoses',<br>'comorbidity - WeightLoss',<br>'comorbidity - Liver',<br>'comorbidity - PHTN',<br>'comorbidity - Drugs',<br>'comorbidity - Mets',<br>'comorbidity - PUD',<br>'comorbidity - HIV' | 'procedure - had cvc',<br>'procedure - had surgery',<br>'procedure - had mechanical ventilation',<br>'procedure - had hemodialysis',<br>'procedure - had parenteral nutrition' | 'infection_sites – RESPIRATORY_TRACT',<br>'infection_sites – BLOOD',<br>'infection_sites – SKIN_SOFTTISSUE',<br>'infection_sites – ABSCESS_OR_FLUID_NOS',<br>'infection_sites – MUCOCUTANEOUS<br>',<br>'infection_sites – GENITOURINARY' | 'selected micro - colonization pressure VAN - higher level',<br>'selected micro - colonization pressure AMC - overall',<br>'selected micro - colonization pressure AMP - overall',<br>'selected micro - colonization pressure ATM - overall',<br>'selected micro - colonization pressure CAZ - overall',<br>'selected micro - colonization pressure CIP - overall',<br>'selected micro - colonization pressure CLI - overall',<br>'selected micro - colonization pressure CRO - overall',<br>'selected micro - colonization pressure DOX - overall',<br>'selected micro - colonization pressure ERY - overall',<br>'selected micro - colonization pressure FEP - overall',<br>'selected micro - colonization pressure FOX - overall',<br>'selected micro - colonization pressure GEN - overall',<br>'selected micro - colonization pressure IPM - overall',<br>'selected micro - colonization pressure LVX - overall',<br>'selected micro - colonization pressure MEM - overall',<br>'selected micro - colonization pressure MIN - overall',<br>'selected micro - colonization pressure MXF - overall',<br>'selected micro - colonization pressure NIT - overall',<br>'selected micro - colonization pressure OXA - overall',<br>'selected micro - colonization pressure PEN - overall',<br>'selected micro - colonization pressure SAM - overall',<br>'selected micro - colonization pressure SXT - overall',<br>'selected micro - colonization pressure TET - overall',<br>'selected micro - colonization pressure TZP - overall',<br>'selected micro - colonization pressure VAN - overall' |

### C Model Hyperparameters and Thresholds:

Table 3: Hyperparameters used for XGBoost, TabNet, and $TabNet^{self}$ models.

| XGBoost | TabNet | $TabNet^{self}$ |
|---|---|---|
| learning_rate=0.1,<br>n_estimators=100,<br>objective='binary:logistic',<br>booster='gbtree',<br>gamma=0,<br>min_child_weight=1,<br>max_delta_step=0,<br>subsample=0.7,<br>colsample_bytree=1,<br>colsample_bylevel=1,<br>colsample_bynode=1,<br>reg_alpha=0,<br>reg_lambda=1,<br>scale_pos_weight=1 | cat_emb_dim:1,<br>optimizer_fn:torch.optim.Adam,<br>learning_rate=2e-2,<br>mask_type:'entmax' | cat_emb_dim:1,<br>optimizer_fn:torch.optim.Adam,<br>learning_rate=1e-1,<br>mask_type:'sparsemax' |

Table 4: Thresholds used to adjust sensitivity/specificity for resistance prediction for NIT, SXT, CIP, and LVX. Thresholds shown for Logistic Regression, TabNet, and XGBoost Models.

| Model | NIT Threshold | SXT Threshold | CIP Threshold | LVX Threshold |
|---|---|---|---|---|
| LR | 0.2041 | 0.2176 | 0.2351 | 0.2131 |
| XGBoost | 0.2161 | 0.2296 | 0.2521 | 0.2416 |
| TabNet | 0.4732 | 0.5198 | 0.4367 | 0.4687 |
| $TabNet^{self}$ | 0.4527 | 0.4442 | 0.4317 | 0.4942 |

### D Feature Importance Results:

Table 5: Top 30 features ranked as most important for predicting resistance to NIT, SXT, CIP, and LVX. Results shown for logistic regression models.

| NIT | | SXT | | CIP | | LVX | |
|---|---|---|---|---|---|---|---|
| Features | Importance | Features | Importance | Features | Importance | Features | Importance |
| micro - prev resistance NIT ALL | 0.16583442 | micro - prev resistance SXT ALL | 0.28034249 | micro - prev resistance CIP ALL | 0.3403487 | micro - prev resistance CIP ALL | 0.30559178 |
| micro - prev resistance NIT 180 | 0.15071255 | micro - prev resistance SXT 180 | 0.25078951 | micro - prev resistance LVX ALL | 0.30660666 | micro - prev resistance LVX ALL | 0.30373326 |
| micro - prev resistance NIT 90 | 0.11945114 | micro - prev resistance SXT 90 | 0.19041384 | ab class 30 - fluoroquinolone | 0.23784769 | ab class 30 - fluoroquinolone | 0.2633915 |
| micro - prev organism Escherichia 180 | 0.08202016 | demographics - is_white | 0.18387394 | micro - prev resistance CIP 180 | 0.23544381 | ab class 90 - fluoroquinolone | 0.2577558 |
| selected micro - colonization pressure AMC 90 - overall | 0.07873466 | ab class 180 - folate_inhibitor | 0.1515269 | ab class 90 - fluoroquinolone | 0.23530273 | ab class 180 - fluoroquinolone | 0.23919102 |
| micro - prev organism Escherichia 90 | 0.07152221 | medication 180 - trimethoprim/sulfamethoxazole | 0.14561013 | ab class 180 - fluoroquinolone | 0.2254336 | micro - prev resistance LVX 180 | 0.2190522 |
| medication 180 - vancomycin | 0.06614874 | ab class 90 - folate_inhibitor | 0.14032988 | micro - prev resistance LVX 180 | 0.21812328 | micro - prev resistance CIP 180 | 0.21028751 |
| ab class 180 - glycopeptides | 0.06614874 | medication 90 - trimethoprim/sulfamethoxazole | 0.13619394 | ab class 14 - fluoroquinolone | 0.17475106 | ab class ALL - fluoroquinolone | 0.19484207 |
| micro - prev organism Klebsiella 180 | 0.06456696 | ab class 30 - folate_inhibitor | 0.12365965 | ab class ALL - fluoroquinolone | 0.17423389 | ab class 14 - fluoroquinolone | 0.19216649 |
| medication ALL - vancomycin | 0.06372871 | ab class ALL - folate_inhibitor | 0.12201817 | micro - prev resistance CIP 90 | 0.17136615 | medication 90 - ciprofloxacin | 0.16934576 |

| | | | | | | | |
|---|---|---|---|---|---|---|---|
| ab class ALL - glycopeptides | 0.06372871 | medication 30 - trimethoprim/sulfamethoxazole | 0.12069678 | micro - prev resistance LVX 90 | 0.16016368 | medication 30 - ciprofloxacin | 0.16871254 |
| medication 90 - vancomycin | 0.06298713 | medication ALL - trimethoprim/sulfamethoxazole | 0.1193667 | medication 90 - ciprofloxacin | 0.15916882 | micro - prev resistance LVX 90 | 0.16210684 |
| ab class 90 - glycopeptides | 0.06298713 | micro - prev resistance CIP ALL | 0.10194257 | medication 30 - ciprofloxacin | 0.15722034 | medication 180 - ciprofloxacin | 0.15781358 |
| micro - prev resistance NIT 30 | 0.05880413 | ab class 14 - folate_inhibitor | 0.09473118 | medication 180 - ciprofloxacin | 0.14927333 | micro - prev resistance CIP 90 | 0.15360967 |
| ab class 30 - glycopeptides | 0.05016528 | medication 14 - trimethoprim/sulfamethoxazole | 0.09218648 | medication ALL - ciprofloxacin | 0.12696075 | medication ALL - levofloxacin | 0.14274374 |
| medication 30 - vancomycin | 0.05016528 | micro - prev resistance LVX ALL | 0.09181497 | medication ALL - levofloxacin | 0.12623817 | medication ALL - ciprofloxacin | 0.13910187 |
| micro - prev organism Klebsiella 90 | 0.049312 | micro - prev resistance GEN ALL | 0.09078446 | micro - prev resistance SXT ALL | 0.12217562 | medication 180 - levofloxacin | 0.13101941 |
| ab class 90 - beta_lactam | 0.0489893 | micro - prev resistance SXT 30 | 0.08348589 | procedure 180 - had surgery | 0.12043989 | medication 90 - levofloxacin | 0.12458092 |
| ab class 30 - beta_lactam | 0.0477703 | micro - prev resistance TET 180 | 0.08194493 | medication 180 - levofloxacin | 0.11898888 | medication 14 - ciprofloxacin | 0.12340371 |
| ab class 180 - beta_lactam | 0.04572374 | micro - prev resistance CIP 180 | 0.07774626 | micro - prev resistance GEN ALL | 0.11660803 | procedure 180 - had surgery | 0.11650756 |
| medication 180 - metronidazole | 0.04214617 | micro - prev resistance TET ALL | 0.07700492 | medication 14 - ciprofloxacin | 0.11543158 | medication 30 - levofloxacin | 0.1133268 |
| ab class 180 - nitroimidazole | 0.04214617 | micro - prev resistance LVX 180 | 0.07663341 | demographics - is_white | 0.10420415 | selected micro - colonization pressure AMC 90 - overall | 0.11095493 |
| micro - prev organism Proteus 180 | 0.04151563 | ab class 30 - fluoroquinolone | 0.07571909 | medication 90 - levofloxacin | 0.10369256 | ab class 7 - fluoroquinolone | 0.11073188 |
| micro - prev resistance AMP 180 | 0.03937871 | micro - prev resistance AMP 180 | 0.07099553 | ab class 7 - fluoroquinolone | 0.1012056 | micro - prev resistance GEN ALL | 0.10628557 |
| selected micro - colonization pressure NIT 90 - overall | 0.03924538 | micro - prev resistance AMP ALL | 0.06838118 | selected micro - colonization pressure AMC 90 - overall | 0.09971343 | micro - prev resistance SXT ALL | 0.1044242 |
| medication 90 - metronidazole | 0.03869241 | micro - prev resistance TET 90 | 0.06365054 | medication ALL - azithromycin | 0.09708152 | medication ALL - azithromycin | 0.09624824 |
| ab class 90 - nitroimidazole | 0.03869241 | ab class 90 - fluoroquinolone | 0.06281437 | medication 30 - levofloxacin | 0.09597308 | ab class ALL - macrolide_lincosamide | 0.09451643 |
| ab class 14 - beta_lactam | 0.03867503 | micro - prev resistance GEN 180 | 0.06224034 | ab subtype ALL - penicillins | 0.09237294 | hosp ward - OP | 0.09238836 |
| micro - prev resistance ATM ALL | 0.03839694 | ab class 180 - fluoroquinolone | 0.06136909 | ab class ALL - macrolide_lincosamide | 0.09127419 | ab subtype ALL - penicillins | 0.08977218 |
| comorbidity 180 - NeuroOther | 0.03774129 | ab class 7 - folate_inhibitor | 0.06111644 | micro - prev resistance SXT 180 | 0.08284649 | selected micro - colonization pressure AMP 90 - overall | 0.08314208 |
| micro - prev resistance CRO ALL | 0.03658179 | medication 7 - trimethoprim/sulfamethoxazole | 0.05970333 | medication ALL - amoxicillin | 0.07912437 | demographics - is_white | 0.08219577 |

Table 6: Top 30 features ranked as most important for predicting resistance to NIT, SXT, CIP, and LVX, alongside importance scores. Results shown for XGBoost models.

| NIT | | SXT | | CIP | | LVX | |
|---|---|---|---|---|---|---|---|
| Features | Importance | Features | Importance | Features | Importance | Features | Importance |
| micro - prev resistance NIT 180 | 0.08351032 | micro - prev resistance SXT 180 | 0.06823729 | micro - prev resistance CIP ALL | 0.08377941 | micro - prev resistance LVX ALL | 0.17187826 |
| ab class 180 - glycopeptides | 0.017065153 | medication 14 - trimethoprim/sulfamethoxazole | 0.01795701 | micro - prev resistance LVX ALL | 0.06061687 | hosp ward - OP | 0.03102657 |
| ab class ALL - glycopeptides | 0.012289353 | ab class 14 - folate_inhibitor | 0.01722852 | hosp ward - OP | 0.03647425 | ab class 30 - fluoroquinolone | 0.02349518 |
| hosp ward - OP | 0.00917343 | medication 30 - trimethoprim/sulfamethoxazole | 0.01527695 | ab class 30 - fluoroquinolone | 0.02084975 | micro - prev resistance CIP 180 | 0.01699777 |
| micro - prev resistance NIT ALL | 0.00748824 | micro - prev resistance SXT ALL | 0.00921823 | micro - prev resistance CIP 180 | 0.01471559 | micro - prev resistance LVX 180 | 0.0125961 |
| micro - prev resistance NIT 30 | 0.004948121 | demographics - is_white | 0.00591628 | micro - prev resistance LVX 180 | 0.01379211 | ab class 90 - fluoroquinolone | 0.00907134 |

| | | | | | | | |
|---|---|---|---|---|---|---|---|
| micro - prev resistance NIT 90 | 0.00414273 | ab class 30 - folate_inhibitor | 0.00577075 | ab class 90 - fluoroquinolone | 0.00746871 | ab class 180 - fluoroquinolone | 0.006308 |
| micro - prev resistance AMK ALL | 0.003790008 | ab class 30 - fluoroquinolone | 0.00487138 | micro - prev resistance CRO 90 | 0.00704285 | micro - prev resistance LVX 90 | 0.00553951 |
| micro - prev resistance TOB 90 | 0.003750979 | micro - prev organism Staph_coag_neg 30 | 0.00474757 | selected micro - colonization pressure LVX 90 - higher level | 0.00523972 | micro - prev resistance CIP ALL | 0.00487915 |
| ab class 90 - glycopeptides | 0.003528108 | ab class 180 - folate_inhibitor | 0.00416789 | ab class 180 - fluoroquinolone | 0.005147 | micro - prev resistance CIP 90 | 0.00432116 |
| medication 30 - cefazolin | 0.003304296 | micro - prev organism Staph_coag_neg 180 | 0.0040985 | micro - prev resistance PEN 90 | 0.00338488 | micro - prev resistance CRO 90 | 0.00403697 |
| comorbidity 30 - Paralysis | 0.002996726 | micro - prev resistance CFZ 180 | 0.00398183 | micro - prev resistance LVX 90 | 0.00323272 | ab class 14 - fluoroquinolone | 0.00354063 |
| ab class 30 - glycopeptides | 0.002804813 | ab subtype 7 - penicillins | 0.0038896 | micro - prev resistance STRS 90 | 0.00321006 | selected micro - colonization pressure LVX 90 - higher level | 0.003357 |
| micro - prev resistance CRO 90 | 0.002613648 | ab class 30 - tetracycline | 0.00384022 | micro - prev resistance ERY 180 | 0.00296634 | micro - prev resistance PEN 90 | 0.00307709 |
| medication 90 - cefoxitin | 0.002595496 | micro - prev resistance SXT 30 | 0.00376002 | custom 90 - nursing home | 0.00293302 | micro - prev resistance ATM 180 | 0.00285963 |
| ab subtype 30 - cephalosporin_gen1 | 0.002531748 | micro - prev organism Staph_coag_neg 90 | 0.00353737 | micro - prev organism Escherichia 180 | 0.00286191 | micro - prev resistance TOB 180 | 0.00280073 |
| custom 90 - nursing home | 0.002494513 | micro - prev resistance PEN 30 | 0.00353185 | micro - prev resistance CIP 90 | 0.00285858 | custom 90 - nursing home | 0.00269507 |
| micro - prev organism Escherichia 180 | 0.002494502 | micro - prev resistance CFZ 90 | 0.00347884 | ab class 14 - fluoroquinolone | 0.00281424 | micro - prev resistance MXF 180 | 0.00261315 |
| ab class ALL - oxazolidinones | 0.002488599 | ab class 90 - folate_inhibitor | 0.00335275 | micro - prev resistance CFZ 90 | 0.00280872 | custom 7 - nursing home | 0.00260861 |
| micro - prev resistance IPM ALL | 0.002478065 | micro - prev resistance SXT 90 | 0.002989 | medication 180 - ertapenem | 0.00273367 | micro - prev resistance FEP ALL | 0.00256069 |
| selected micro - colonization pressure LVX 90 - higher level | 0.002436119 | comorbidity 30 - HIV | 0.00291577 | comorbidity 7 - Paralysis | 0.00261325 | ab class ALL - fluoroquinolone | 0.00242046 |
| micro - prev resistance ATM ALL | 0.002429983 | micro - prev resistance CIP 180 | 0.00288673 | micro - prev resistance FEP ALL | 0.00253271 | micro - prev resistance ERY 180 | 0.00237031 |
| comorbidity 180 - FluidsLytes | 0.002419386 | micro - prev resistance CRO 30 | 0.00286796 | selected micro - colonization pressure CIP 90 - higher level | 0.0025202 | ab subtype 90 - cephalosporin_gen3 | 0.00235223 |
| micro - prev organism Proteus 90 | 0.002360134 | micro - prev resistance CFZ ALL | 0.00285349 | micro - prev resistance ATM 180 | 0.00251334 | micro - prev resistance CFZ 180 | 0.00234603 |
| medication 30 - dicloxacillin | 0.002357808 | ab subtype 14 - penicillins | 0.0028375 | ab class ALL - fluoroquinolone | 0.00243381 | comorbidity 14 - Paralysis | 0.00232654 |
| ab subtype 30 - antistaphylococcal | 0.002355573 | medication 90 - trimethoprim/sulfa methoxazole | 0.00282847 | ab subtype ALL - carbapenem | 0.00233752 | medication 90 - dicloxacillin | 0.00222465 |
| micro - prev organism Escherichia 90 | 0.002331048 | micro - prev resistance AMP 90 | 0.00276171 | micro - prev resistance CFZ 180 | 0.00232454 | ab subtype 14 - cephalosporin_gen3 | 0.0022049 |
| comorbidity 180 - DM | 0.002328892 | micro - prev organism Escherichia 90 | 0.0027339 | comorbidity 7 - WeightLoss | 0.00222559 | comorbidity 7 - Paralysis | 0.0021903 |
| ab class 7 - nitrofuran | 0.002324742 | ab class 14 - fluoroquinolone | 0.00271177 | micro - prev resistance SAM 180 | 0.0021894 | micro - prev organism Staph_coag_neg 90 | 0.00218047 |
| ab subtype 14 - cephalosporin_gen1 | 0.002320632 | micro - prev organism Escherichia 180 | 0.00264852 | ab subtype 14 - cephalosporin_gen1 | 0.00215853 | medication 180 - ampicillin/sulbactam | 0.002085 |
| comorbidity 14 - Renal | 0.002294314 | custom 90 - nursing home | 0.00259581 | micro - prev resistance ATM ALL | 0.00213543 | medication 180 - cefpodoxime | 0.00208406 |

Table 7: Top 30 features ranked as most important for predicting resistance to NIT, SXT, CIP, and LVX. Results shown for TabNet models.

| NIT | | SXT | | CIP | | LVX | |
|---|---|---|---|---|---|---|---|
| Features | Importance | Features | Importance | Features | Importance | Features | Importance |
| micro - prev resistance NIT 180 | 0.14030591 | micro - prev resistance SXT 90 | 0.18105172 | micro - prev resistance CIP ALL | 0.13274792 | micro - prev resistance CIP ALL | 0.14050733 |

| | | | | | | | |
|---|---|---|---|---|---|---|---|
| selected micro - colonization pressure IPM 90 - overall | 0.12083684 | micro - prev resistance OXA 180 | 0.16303417 | ab subtype 14 - antistaphylococcal | 0.12223279 | ab class 30 - fluoroquinolone | 0.100545319 |
| micro - prev resistance NIT 90 | 0.09461148 | selected micro - colonization pressure PEN 90 - granular level | 0.1113221 | ab class 90 - fluoroquinolone | 0.10249452 | micro - prev resistance LVX 90 | 0.090840053 |
| micro - prev resistance NIT ALL | 0.05946949 | comorbidity 30 - Obesity | 0.09189122 | micro - prev resistance CIP 180 | 0.09147021 | ab class 90 - fluoroquinolone | 0.074081709 |
| micro - prev resistance CFZ 30 | 0.05284526 | medication 14 - trimethoprim/sulfamethoxazole | 0.07772281 | hosp ward - OP | 0.08934388 | micro - prev resistance CIP 180 | 0.056748595 |
| medication 90 - vancomycin | 0.05268151 | ab class 30 - folate_inhibitor | 0.04816563 | ab class 30 - fluoroquinolone | 0.06785838 | hosp ward - OP | 0.055764986 |
| comorbidity 7 - CHF | 0.052296 | comorbidity 180 - HTN | 0.04219703 | micro - prev resistance LVX 180 | 0.05404586 | ab class 14 - fluoroquinolone | 0.047868894 |
| micro - prev resistance NIT 30 | 0.05171871 | micro - prev resistance CIP 90 | 0.0297893 | ab class 180 - glycopeptides | 0.04559108 | micro - prev resistance LVX 180 | 0.044784657 |
| medication 30 - vancomycin | 0.03474571 | ab class 30 - fluoroquinolone | 0.02746249 | ab subtype ALL - cephalosporin_gen4 | 0.03724927 | medication 180 - ceftriaxone | 0.044292283 |
| ab class 180 - nitroimidazole | 0.03127979 | comorbidity 7 - HTNcx | 0.0252487 | micro - prev resistance LVX 90 | 0.02698228 | medication 180 - levofloxacin | 0.038854445 |
| comorbidity 180 - Paralysis | 0.03112449 | comorbidity 30 - PVD | 0.02395814 | micro - prev resistance TOB 30 | 0.02676638 | custom 14 - nursing home | 0.038688439 |
| ab class 14 - nitrofuran | 0.03025161 | micro - prev resistance CIP ALL | 0.01686783 | custom 30 - nursing home | 0.02071371 | medication 180 - cefepime | 0.032634041 |
| medication ALL - cefepime | 0.02678745 | ab class 180 - fluoroquinolone | 0.01567822 | selected micro - colonization pressure AMP 90 - higher level | 0.01801438 | micro - prev resistance FEP 180 | 0.027690859 |
| comorbidity 14 - FluidsLytes | 0.02363112 | medication 14 - erythromycin | 0.01517378 | micro - prev organism Proteus 30 | 0.01780917 | micro - prev resistance LVX ALL | 0.024022646 |
| comorbidity 180 - HTNcx | 0.02106431 | medication ALL - trimethoprim/sulfamethoxazole | 0.01489986 | micro - prev resistance NIT 14 | 0.01712635 | ab class ALL - fluoroquinolone | 0.01846237 |
| ab subtype 180 - cephalosporin_gen4 | 0.0207901 | micro - prev resistance LVX 180 | 0.01383609 | comorbidity 180 - BloodLoss | 0.01522945 | medication 7 - erythromycin | 0.017251008 |
| medication ALL - metronidazole | 0.01977575 | ab class 180 - antifungal | 0.01166533 | comorbidity 180 - Liver | 0.01317582 | ab class 180 - fluoroquinolone | 0.016300964 |
| ab class 30 - beta_lactam | 0.01896005 | ab class 7 - folate_inhibitor | 0.01095832 | micro - prev resistance SAM 30 | 0.01170047 | comorbidity 90 - Paralysis | 0.016045939 |
| ab class 90 - fosfomycin | 0.01392381 | comorbidity 7 - DM | 0.00943496 | medication 7 - ceftazidime | 0.0103175 | micro - prev organism Escherichia 180 | 0.01434185 |
| medication 180 - meropenem | 0.01147187 | medication 30 - clarithromycin | 0.00923156 | medication 30 - cephalexin | 0.00989095 | micro - prev resistance ATM 90 | 0.01147941 |
| selected micro - colonization pressure AMP 90 - higher level | 0.01080954 | micro - prev resistance TET 180 | 0.00545259 | comorbidity 14 - Mets | 0.007243 | micro - prev organism Escherichia 14 | 0.0096465 |
| ab class 7 - antifungal | 0.00836307 | selected micro - colonization pressure AMC 90 - higher level | 0.00350792 | medication 180 - linezolid | 0.00678098 | micro - prev resistance GEN 30 | 0.007449921 |
| infection_sites - RESPIRATORY_TRACT | 0.00600549 | medication 30 - rifampin | 0.00332839 | micro - prev resistance TET 30 | 0.00459549 | comorbidity 90 - CHF | 0.006721011 |
| medication 90 - nafcillin | 0.00539483 | ab class ALL - mixed | 0.0028207 | medication 180 - cephalexin | 0.00420383 | micro - prev resistance IPM ALL | 0.005981251 |
| comorbidity 14 - NeuroOther | 0.00506691 | micro - prev organism Enterobacter 30 | 0.0027973 | micro - prev organism Staph_coag_neg 30 | 0.00362137 | comorbidity 90 - Renal | 0.005589539 |
| ab class 7 - fluoroquinolone | 0.00487837 | comorbidity 90 - HTNcx | 0.00269018 | ab class 7 - glycopeptides | 0.00358605 | micro - prev resistance IPM 180 | 0.005234392 |
| medication 180 - vancomycin | 0.00478214 | infection_sites - BLOOD | 0.00215572 | ab class 14 - fluoroquinolone | 0.00357785 | selected micro - colonization pressure SXT 90 - granular level | 0.004104467 |
| micro - prev resistance NAL ALL | 0.00440548 | comorbidity 180 - HIV | 0.00207932 | medication 180 - vancomycin | 0.0035629 | medication ALL - levofloxacin | 0.003853533 |
| comorbidity 7 - Obesity | 0.00432116 | comorbidity 180 - Paralysis | 0.00202259 | ab class 14 - beta_lactam | 0.00235753 | comorbidity 30 - Arrhythmia | 0.00352799 |
| micro - prev resistance NIT 14 | 0.00314751 | medication 180 - amikacin | 0.00198349 | medication 30 - ceftriaxone | 0.00196161 | ab subtype 14 - cephalosporin_gen2 | 0.00334722 |

| | | | | | | | |
|---|---|---|---|---|---|---|---|
| ab subtype 14 - antistaphylococ cal | 0.0030716 | micro - prev resistance DOX 180 | 0.00191412 | comorbidity 7 - Anemia | 0.00185372 | comorbidity 14 - Renal | 0.002813001 |

Table 8: Top 30 features ranked as most important for predicting resistance to NIT, SXT, CIP, and LVX. Results shown for TabNet$^{self}$ models.

| NIT | | SXT | | CIP | | LVX | |
|---|---|---|---|---|---|---|---|
| Features | Importance | Features | Importance | Features | Importance | Features | Importance |
| micro - prev resistance TZP 90 | 0.38602463 | micro - prev resistance SXT 90 | 0.18960561 | micro - prev resistance QUD 30 | 0.20761771 | medication 90 - metronidazole | 0.43875042 |
| custom 90 - nursing home | 0.06491553 | selected micro - colonization pressure CAZ 90 - overall | 0.12493466 | micro - prev organism Proteus 90 | 0.08499887 | comorbidity 90 - Paralysis | 0.15263563 |
| micro - prev organism Klebsiella 30 | 0.0596158 | medication 90 - ampicillin/sulbactam | 0.09458135 | ab subtype 180 - carbapenem | 0.07866491 | medication 7 - amoxicillin | 0.05811898 |
| ab subtype 90 - antistaphylococcal | 0.0585334 | micro - prev resistance SXT ALL | 0.08413758 | medication 180 - clindamycin | 0.06356454 | medication 7 - levofloxacin | 0.05783063 |
| medication 180 - ofloxacin | 0.04245956 | medication 90 - ceftazidime | 0.08319119 | micro - prev resistance CIP ALL | 0.05239772 | comorbidity 14 - Mets | 0.04136833 |
| micro - prev resistance NIT 90 | 0.03451244 | micro - prev resistance TET 180 | 0.06189176 | micro - prev resistance CIP 90 | 0.03945457 | micro - prev resistance AMC 180 | 0.0272212 |
| micro - prev resistance GEN 90 | 0.02481275 | micro - prev resistance SXT 180 | 0.04004747 | micro - prev resistance CIP 180 | 0.03758965 | infection_sites - GENITOURINARY | 0.02167372 |
| comorbidity 14 - Hypothyroid | 0.02389192 | micro - prev organism Staph_coag_neg 90 | 0.03231319 | micro - prev resistance PIP 180 | 0.0360328 | ab class ALL - fluoroquinolone | 0.01979572 |
| micro - prev resistance NIT 30 | 0.02318466 | medication ALL - meropenem | 0.02801294 | medication 14 - dicloxacillin | 0.03311215 | ab class ALL - antifungal | 0.01571733 |
| micro - prev resistance NIT 14 | 0.02098432 | micro - prev resistance GEN 180 | 0.02798877 | medication 90 - ciprofloxacin | 0.02847176 | micro - prev resistance ERY 90 | 0.01531765 |
| ab class 180 - glycopeptides | 0.02083964 | micro - prev resistance CIP 90 | 0.02450381 | medication 30 - levofloxacin | 0.02833882 | micro - prev resistance CHL 90 | 0.01361284 |
| micro - prev resistance NIT 180 | 0.01752002 | medication 90 - ertapenem | 0.02435632 | micro - prev resistance LVX 180 | 0.02595891 | ab subtype 180 - cephalosporin_gen3 | 0.01049224 |
| medication 90 - dicloxacillin | 0.01740196 | comorbidity 90 - Pulmonary | 0.02275696 | medication 30 - penicillin | 0.01828632 | micro - prev organism Acinetobacter 180 | 0.01008098 |
| medication 180 - amphotericin_b | 0.01347599 | micro - prev organism Acinetobacter 90 | 0.01707451 | medication 180 - fluconazole | 0.01763353 | micro - prev resistance LVX 180 | 0.00796976 |
| ab subtype 90 - cephalosporin_gen2 | 0.01133894 | medication 180 - cefoxitin | 0.01180612 | micro - prev resistance LVX ALL | 0.01594754 | ab subtype 14 - penicillins | 0.00726561 |
| ab subtype 180 - azole | 0.01108682 | micro - prev resistance CLI ALL | 0.01007763 | ab subtype 30 - antistaphylococcal | 0.01541937 | micro - prev resistance NAL 180 | 0.00721123 |
| ab subtype 14 - azole | 0.0110121 | ab class ALL - nitroimidazole | 0.01001294 | micro - prev resistance AMP ALL | 0.01383369 | medication ALL - metronidazole | 0.00613392 |
| medication 90 - rifampin | 0.01090429 | micro - prev resistance LVX 14 | 0.00914347 | comorbidity 180 - Renal | 0.01207362 | micro - prev resistance AMK 90 | 0.00540971 |
| selected micro - colonization pressure CRO 90 - granular level | 0.00968358 | medication 90 - tetracycline | 0.00553797 | medication 180 - metronidazole | 0.01116693 | ab class 30 - aminoglycoside | 0.0050919 |
| medication 180 - amoxicillin | 0.00937788 | comorbidity 180 - DMcx | 0.00539228 | comorbidity 30 - HTN | 0.01081988 | micro - prev resistance GEN 90 | 0.00491015 |
| micro - prev organism Serratia 180 | 0.00918053 | micro - prev resistance AMP 90 | 0.00535982 | medication 90 - fluconazole | 0.01002798 | micro - prev organism Streptococcus 90 | 0.0045545 |
| custom 30 - nursing home | 0.00777287 | ab class 180 - monobactam | 0.0050586 | micro - prev resistance IPM ALL | 0.00945698 | micro - prev resistance QUD 180 | 0.00430664 |
| micro - prev resistance CIP 180 | 0.00738211 | medication 30 - doxycycline | 0.00495242 | micro - prev organism Streptococcus 30 | 0.00895859 | micro - prev resistance ERY ALL | 0.0041718 |
| ab subtype ALL - antistaphylococcal | 0.00728373 | comorbidity 14 - FluidsLytes | 0.00495142 | medication 7 - cefadroxil | 0.00801943 | medication 7 - cefpodoxime | 0.00403025 |
| micro - prev organism Enterococcus 180 | 0.00482161 | ab class ALL - nitrofuran | 0.00445599 | medication 180 - cefprozil | 0.00772892 | micro - prev resistance AMP 90 | 0.00345208 |
| comorbidity 7 - Hypothyroid | 0.00479794 | micro - prev resistance ATM ALL | 0.00439387 | selected micro - colonization pressure SXT 90 - higher level | 0.00736957 | micro - prev resistance CTT 180 | 0.00303039 |
| medication 180 - moxifloxacin | 0.00468081 | comorbidity 7 - HTNcx | 0.00420613 | ab class 90 - fosfomycin | 0.00671697 | ab class ALL - ansamycin | 0.00239813 |

| | | | | | | | |
|---|---|---|---|---|---|---|---|
| medication ALL - linezolid | 0.00432525 | medication 180 - metronidazole | 0.00397263 | selected micro - colonization pressure MEM 90 - higher level | 0.0062214 | medication 90 - cefepime | 0.0022582 |
| micro - prev resistance CHL 180 | 0.00388634 | micro - prev resistance DOX 30 | 0.0039153 | micro - prev resistance NIT 180 | 0.0058609 | comorbidity 14 - Psychoses | 0.00212862 |
| ab class 7 - antifungal | 0.00388363 | comorbidity 90 - Coagulopathy | 0.00343889 | ab class 14 - ansamycin | 0.00499642 | medication 180 - trimethoprim | 0.00205345 |
| micro - prev resistance NIT ALL | 0.00386439 | medication ALL - trimethoprim | 0.00309072 | micro - prev resistance CAZ ALL | 0.00492482 | micro - prev organism Enterococcus 180 | 0.00194181 |

### E Additional Results:

Table 9: AUROC and AUPRC scores of XGBoost models trained without particular feature sets, with error bars representing 95% CIs.

| | NIT | | SXT | | CIP | | LVX | |
|---|---|---|---|---|---|---|---|---|
| | AUROC | AUPRC | AUROC | AUPRC | AUROC | AUPRC | AUROC | AUPRC |
| Prior antibiotic exposure | 0.677 (0.671-0.683) | 0.402 (0.393-0.412) | 0.671 (0.665-0.677) | 0.484 (0.474-0.494) | 0.784 (0.779-0.789) | 0.579 (0.569-0.589) | 0.787 (0.783-0.792) | 0.582 (0.573-0.593) |
| Prior antibiotic resistance | 0.674 (0.668-0.680) | 0.391 (0.382-0.402) | 0.649 (0.643-0.656) | 0.437 (0.427-0.447) | 0.771 (0.765-0.776) | 0.548 (0.537-0.558) | 0.773 (0.768-0.778) | 0.561 (0.551-0.571) |
| Prior infecting organisms | 0.682 (0.676-0.688) | 0.404 (0.393-0.414) | 0.701 (0.695-0.707) | 0.523 (0.512-0.533) | 0.808 (0.803-0.813) | 0.613 (0.604-0.622) | 0.813 (0.808-0.818) | 0.620 (0.610-0.630) |

Table 10: AUPRC score decreases of XGBoost models trained without particular feature sets compared to XGBoost models trained with all feature sets, with error bars representing 95% CIs. P-values calculated using 1000 bootstrap samples comparing AUPRC difference.

| | NIT | | SXT | | CIP | | LVX | |
|---|---|---|---|---|---|---|---|---|
| | AUPRC | p-value | AUPRC | p-value | AUPRC | p-value | AUPRC | p-value |
| Prior antibiotic exposure | 0.0089 (0.0035-0.0142) | 0.001 | 0.0401 (0.0035-0.0142) | <0.001 | 0.0383 (0.0327-0.0441) | <0.001 | 0.0414 (0.0352-0.0472) | <0.001 |
| Prior antibiotic resistance | 0.0199 (0.0142- 0.0265) | <0.001 | 0.0877 (0.0800-0.0947) | <0.001 | 0.0695 (0.0623-0.0759) | <0.001 | 0.0631 (0.0566-0.0692) | <0.001 |
| Prior infecting organisms | 0.0049 (0.0019-0.0072) | 0.001 | 0.0015 (-0.0019-0.0052) | 0.403 | 0.0038 (0.000-0.0079) | 0.059 | 0.0034 (-0.0003-0.0068) | 0.062 |

Table 11: AUROC score decreases of XGBoost models trained without particular feature sets compared to XGBoost models trained with all feature sets, with error bars representing 95% CIs. P-values calculated using 1000 bootstrap samples comparing AUROC difference.

| | NIT | | SXT | | CIP | | LVX | |
|---|---|---|---|---|---|---|---|---|
| | AUROC | p-value | AUROC | p-value | AUROC | p-value | AUROC | p-value |
| Prior antibiotic exposure | 0.0093 (0.0068-0.0130) | <0.001 | 0.0302 (0.0262-0.0343) | <0.001 | 0.0269 (0.0243-0.0296) | <0.001 | 0.0270 (0.0243-0.0298) | <0.001 |
| Prior antibiotic resistance | 0.0126 (0.0089- 0.0166) | <0.001 | 0.0520 (0.0472-0.0569) | <0.001 | 0.0405 (0.0373-0.0435) | <0.001 | 0.0411 (0.0382-0.0442) | <0.001 |
| Prior infecting organisms | 0.0077 (0.0029-0.0123) | 0.001 | 0.0002 (-0.0024-0.0027) | 0.910 | 0.0030 (0.0015-0.0045) | <0.001 | 0.0015 (0.0003-0.0026) | 0.011 |
